# Vitamin D levels and COVID-19 severe pneumonia: a prospective case-control study

**DOI:** 10.1101/2024.06.10.24308690

**Authors:** Fortunato José Cardoso, Carla Adriane Leal de Araújo, José Roberto da Silva Junior, Angélica Guimarães, Michelle Viana Taveiro, José Natal Figueiroa, João Guilherme Bezerra Alves

## Abstract

**Introduction:** The hypothesis that a low vitamin D levels is associated with a higher risk for severe COVID-19 has not been completely proven, especially with severe pneumonia.

**Objective:** The goal of this study was to confirm the link between vitamin D levels and COVID-19 severe pneumonia.

**Methods:** This prospective case-control study involved 307 patients who developed severe SARS-CoV-2 pneumonia and were hospitalized in an intensive care unit. Age- and sex-matched controls (307) were selected from the same population; 307 patients with mild to moderate forms of COVID-19 who were not hospitalized. Vitamin D levels were assessed during the duration of the disease.

**Results:** The mean vitamin D level was lower in the severe COVID-19 pneumonia group as compared to the control group; 26.8 ± 7.6 ng/mL *vs* 28.6 ± 7.4 ng/mL, p<0.002. There were more patients with a sufficient level of vitamin D in the control group as compared to the control group; 127 (20.6%) vs 89 (14.5%), p<0.001. Multivariable analysis showed that a deficient vitamin D level was associated with a higher risk for severe COVID-19 pneumonia (OR=3.0; 95% CI: 1.79, 5.10CI), p<0.001.

**Conclusion:** A sufficient vitamin D level is linked to a lower risk of COVID-19 severe pneumonia.

## Introduction

The COVID-19 pandemic has become a global threat, not only because of its impact on health but also because of the great economic burden it has placed on the lives of the affected populations [1]. Although in the last few months the pandemic has decreased worldwide, a new wave of COVID has arisen in the last few months in some countries with SARS-CoV-2 Omicron subvariants, especially BQ.1, BQ.1, and XBB [2, 3].

Infection with SARS-CoV-2 is silent or a benign upper respiratory disease in 80% of cases and causes pneumonia in 20% of cases [4, 5]. Approximately half of these patients develop hypoxemic pneumonia, requiring hospitalization and, sometimes, UTI admission and mechanical ventilation. Pneumonia represents the typical presentation of COVID-19 and is the main mortality cause [6, 7]. SARS-CoV-2 pneumonia can lead to acute hypoxic respiratory failure, multiorgan failure, and death [8, 9]. SARS-CoV-2 leads to diffuse alveolar damage and micro- and macrothrombi in pulmonary arterial vessels, resulting in profound oxygen desaturation and respiratory distress [10].

Due to the presence of the vitamin D receptor in various types of cells and tissues, vitamin D has many biologic activities in various organ systems and acts as an immunomodulatory factor in the prevention of respiratory infections [11, 12]. Some studies have reported that vitamin D may be involved both in the SARSCoV-2 replication process and in the binding of the virus to the host [13, 14, 15]. COVID-19 has been more commonly reported in regions with populations with low vitamin D, including patients with severe COVID-19 disease [16, 17, 18, 19]. However, these studies have demonstrated conflicting results. A recent meta-analysis showed that the evidence of vitamin D protection against hospitalization, ICU admission, and pulmonary involvement is currently inconsistent and insufficient [20]. Another recent systematic review concluded that deficient vitamin D levels were not associated with an increased mortality rate in patients with COVID-19 when the analysis included studies with adjustments for confounders [21]. In conclusion, the relationship between COVID-19 and vitamin D has biological plausibility, but the studies available currently are not conclusive [22].

The hypothesis that a low vitamin D levels is associated with a higher risk for severe COVID-19 has not been completely proven, especially with severe pneumonia. Based on all this, this case-control study verified the association between vitamin D level and the risk of severe COVID-19 pneumonia.

## Subjects and Methods

This prospective matched case-control study was performed in Recife, Northeast, Brazil. Patients were enrolled at the Instituto de Medicina Integral Prof. Fernando Figueira (IMIP), and Hospital Dom Malan (HDM). Case and control patients were selected between April 2020 and February 2022 from patients treated with COVID-19 at IMIP and HDM.

This observational study followed the ethical standards of the 1964 Declaration of Helsinki and its later amendments. This study was previously approved by the Research Ethical Committee of IMIP (CAAE: 40857920.0.0000.5201). Written informed consent was previously obtained from each participant.

Patients hospitalized in an intensive care unit with COVID-19 severe pneumonia and over the age of 18 who were considered case patients. COVID-19 severe pneumonia was considered according to the following definition: shortness of breath, low SpO2, and pulmonary alteration on a CT image. A positive reverse transcription polymerase chain reaction (RT-PCR) test of pharyngeal and nasal swab samples was used to confirm the SARS-Cov-2 infection. For every case patient, a patient with mild or moderate COVID-19 was selected as a control, always matched by age (±5 years) and gender. Controls were selected from the same hospitals.

The vitamin D level was assessed using a chemiluminescence-based immunoassay analyzer. Pacients were classified according to vitamin D values as deficient (<20 ng/mL), insufficient (≥ 20 and ≤ 30 ng/mL), and sufficient (>30 ng/mL).

The sample size was determined based on the following information: (1) significance level of 0.05; (2) power=0.8; (3) proportion of those who were exposed to the protection factor (sufficient vitamin D level) versus those who were not exposed (deficient or insufficient vitamin D level); a difference of 12% in the outcome (COVID-19 severe pneumonia); the value of the minimum odds ratio to be detected (0.2); and a case-to-control ratio of one case to one control. A total of 614 patients were calculated;

Categorical variables are presented as frequencies and percentages, while continuous variables are presented as means and standard deviations (SDs). The demographic and clinical variables of the case and control patients were compared using the chi-square test or Student’s t-test. Multivariable logistic regression was used to assess the independent association between vitamin D levels and SARS-CoV-2 severe pneumonia after adjustment for categorical and continuous variables. Results were expressed as odds ratios (ORs) with 95% confidence intervals (CIs). Statistical analyses were performed using STATA version 12.1 (USA). A p value of 0.05 was considered statistically significant.

## Results

A total of 314 patients were studied; 307 case patients hospitalized with COVID-19 severe pneumonia, and 307 matched controls for age and gender, all PCR positive for SARS-CoV-2. There was no difference in the mean age (49.2 ± 13.0 years vs. 48.7 ± 11.0 years; p = 0.631) or gender (307 males vs. 307 females), respectively, in the cases or control patients. Obesity, overweight, systemic hypertension, type 2 diabetes, smoking, and drinking habits showed no differences between cases and controls (Table 1).

**Table 1.** Some characteristics between case and control groups.

Variable Case (307) Control (307) p
|  |  |  |  |
| --- | --- | --- | --- |
| Obesity | 41 (13.3%) | 27 (8.7%) | 0.071 |
| Overweight | 179 (58.3%) | 158 (51.5%) | 0.089 |
| Smoking | 23 (7.4%) | 19 (6.1%) | 0.523 |
| Drinking | 146 (47.6%) | 142 (46.2%) | 0.746 |
| Diabetes Type 2 | 44 (14.3%) | 38 (12.3%) | 0.477 |
| Hypertension | 110 (35.8%) | 117 (38.1%) | 0.558 |

The mean vitamin D level was lower in case patients as compared to control patients, respectively: 26.8 ± 7.6 ng/mL *vs.* 28.6 ± 7.4 ng/mL, p < 0.002. Vitamin D deficiency and insufficiency were more common in the case group (p < 0.001) (Table 2). Multivariable analysis showed that a low vitamin D level was independently associated with a higher risk for SARS-CoV-2 severe pneumonia, 95% CI (1.38 – 3.01), p < 0.001 (Table 3).

**Table 2.** Univariable analysis of some variables between patients with (Cases) and without (Controls) severe SARS-CoV-19 pneumonia.

| Variable | Sample | Case | Control | OR (IC95%) | Valor p |
| --- | --- | --- | --- | --- | --- |
|  | N | (N = 307)<br>N (%) | (N = 307)<br>N (%) |  |  |
| **Diabetes type 2** |  |  |  |  | 0.480 |
| Yes | 82 | 44 (14.3) | 38 (12.4) | 1.2 (0.74 - 1.88) |  |
| No | 532 | 263 (85.7) | 269 (87.6) | 1.0 |  |
| **Smoking** |  |  |  |  | 0.538 |
| Yes | 42 | 23 (7.5) | 19 (6.2) | 1.2 (0.66 - 2.22) |  |
| No | 572 | 284 (92.5) | 288 (93.8) | 1.0 |  |
| **Drinking** |  |  |  |  | 0.742 |
| Yes | 288 | 146 (47.6) | 142 (46.2) | 1.1 (0.76 - 1.46) |  |
| No | 326 | 161 (52.4) | 165 (53.8) | 1.0 |  |
| **Hipertension** |  |  |  |  | 0.566 |
| Yes | 227 | 109 (34.5) | 118 (38.4) | 0.9 (0.66 - 1.26) |  |
| No | 387 | 198 (64.5) | 189 (61.6) | 1.0 |  |
| **Nutritional Status** |  |  |  |  | 0.0971 |
| Normal | 216 | 80 (26.1) | 136 (44.3) | 1.0 |  |
| Overweight | 337 | 179 (58.3) | 158 (51.5) | 1.9 (1.33 – 2.68) |  |
| Obesity | 68 | 41 (13.3) | 27 (8.7) | 3.4 (3.14 – 8.90) |  |
| **Vitamin D level** |  |  |  |  | < 0.001 |
| Insuficient | 294 | 162 (55,1) | 132 (44,9) | 2.2 (1.51 – 3.45) | < 0.001 |
| Deficient | 90 | 62 (68,9) | 28 (31,1) | 3.4 (2.08 – 5.75) | < 0.001 |
| Suficient | 230 | 83 (36,1) | 147 (63,9) | 1.0 |  |

**Table 3.** Multivariable analysis of some variables between patients with (Cases) and without (Controls) severe SARS-CoV-19 pneumonia.

| Variable | Multivariable Model |  |
| --- | --- | --- |
|  | OR (95% CI) | p value |
| **Nutritional Status** |  |  |
| Normal | 1.0 | 0.093 |
| Overweight | 1.8 (1.26 – 2.61) |  |
| Obesity | 3.3 (2.84 – 8.34) |  |
| **Vitamin D Level** |  | <0.001 |
| Insufficient | 2.0 (1.38 – 3.01) |  |
| Deficient | 3.0 (1.79 – 5.10) |  |
| Sufficient | 1.0 |  |

## Discussion

This prospective case-control study showed that a sufficient vitamin D level was associated with a lower risk of SARS-CoV-19 severe pneumonia. Inversely, a deficient or insufficient vitamin D level increased the risk of SARS-CoV-2-related severe pneumonia. Our findings are in agreement with other observational studies that have approached the severity of illness in COVID-19. However, unlike other studies, our severity criterion used in this study was that of severe pneumonia, i.e., admission to an intensive care unit and complaints of shortness of breath, low SpO2, and pulmonary alteration on a CT image. Nimavat et al. found no statistical difference between the mean vitamin D level among 156 cases and 204 controls (p = 0.757), but vitamin D deficiency was observed in 37.1 % of severe pneumonia cases compared to 20 % and 10.1 % among mild and moderate cases, respectively [23]. However, only 23 patients were studied with COVID-19 severe pneumonia. Ye et al. also reported a higher percentage of vitamin D deficiency among patients with severe disease as compared to mild/moderate disease, but they studied only 10 patients with COVID-19 severe pneumonia [24]. Israel et al. in a large retrospective case-control study based on an Israeli database, showed a significant association between vitamin D deficiency and the risks of SARS-CoV-2 severe disease [25]. However, they measured vitamin D levels a long time before hospitalization. These findings, however, are not unanimous. Novakovic et al. studied 685 patients hospitalized with COVID-19 and did not find an association between vitamin D level and 30-day mortality [26].

Some systematic reviews have also examined the association between vitamin D levels and COVID-19 severity. These reviews tend to converge on an inverse association between vitamin D levels and COVID-19 severity. However, the authors emphasize the need for further studies because of the low or moderate quality of the studies. Pereira et al. [27] concluded that vitamin D deficiency was not associated with a higher chance of infection by COVID-19 (OR = 1.35; 95% CI = 0.80-1.88), but that severe cases of COVID-19 present 64% (OR = 1.64; 95% CI = 1.30-2.09) more vitamin D deficiency compared with mild cases. Ebrahimzadeh et al. [28] found a significant direct association between vitamin D deficiency and an elevated risk of COVID-19 in-hospital mortality. Kazemi et al. [29] concluded that lower vitamin D levels correlated with disease severity and a poor prognosis, but most of the studies were observational and of moderate quality. On the other hand, Al Kiyumi et al. [30] concluded that the evidence of vitamin D deficiency and COVID-19 hospitalization, ICU admission, and pulmonary involvement, was inconsistent and insuficient.

It has been hypothesized that there are some potential mechanisms to explain the observed association between vitamin D levels and SARS-CoV-2 infection severity. Vitamin D competes to maintain cell junctions and has protective effects against thrombosis, endothelial dysfunction, and thrombosis. SARS-CoV-2 disrupts cell junction integrity [31]. Vitamin D also enhances cellular innate immunity and interferes with viral replication through the induction of antimicrobial peptides [32]. Vitamin D still promotes the production of antimicrobial and antiviral proteins (β-defensin 2 and cathelicidin), both of which inhibit the replication of SARS-CoV-2 and stimulate the clearance of virus from cells by autophagy [32]. Besides, it has been suggested that vitamin D stimulates ACE2 receptors, which can bind to SARS-CoV-2 and prevent it from attaching to ACE2 receptors and entering the cells [33]. Finally, vitamin D has anti-inflammatory effects, including inhibition of TNF-α (tumor necrosis factor-α) and IL-6 which can help to avoid a cytokine storm and hyper-inflammatory state caused by SARS-CoV-2 and associated with high mortality [33].

Our research has strengths and limitations. This case control study had prospective recruitment and contemporaneous control subjects, and only patients with COVID-19 severe pneumonia with intensive care admission were included. We did not identify observational studies exploring this association in tropical regions where people are exposed to sunlight almost all days of the year. Besides, our study included two health reference centers in Pernambuco State, Brazil, and had the sample size calculated. However, our findings must be interpreted with caution because of certain limitations. First, we could not control the vaccine response in case studies or control patients because, until the middle of the study, vaccines for COVID were not yet being applied in Brazil. Second, we did not include cases of death. Our study design included patients in the intensive care unit, and they had to be able to respond to a questionnaire at the time of discharge from the intensive care unit, which made it impossible to study patients who died.

This finding emphasizes the protective effects of a sufficient vitamin D level against the risks of COVID-19 severe pneumonia. This result has critical implications for public health, especially considering that, until now, COVID-19 had no specific treatment. Thus, it is mandatory to encourage the general population to keep their vitamin D levels sufficient in their everyday routine to reduce the risk of COVID-19 severe pneumonia, the main cause of COVID-19 mortality.

## Conclusion

Previous studies have shown that vitamin D levels are associated with the severity of illness and mortality in COVID-19 patients. This study adds that a sufficient vitamin D level offers protection against COVID-19 severe pneumonia hospitalization and admission to the intensive care unit. Further clinical trial studies are needed to confirm these findings of the beneficial effect of vitamin D on COVID-19 severe pneumonia.

## Data Availability

All data produced in the present study are available upon reasonable request to the authors

## Ethics approval and consent to participate

This study followed the ethical standards of the 1964 Declaration of Helsinki and its later amendments. Written informed consent was obtained previously from each participant. This study was previously approved by the Research Ethical Committee of Instituto de Medicina Integral Prof. Fernando Figueira (IMIP); CAAE (Certificate of Presentation of Ethical Appreciation): 40857920.0.0000.5201.

## Availability of data and materials

Data are available from the authors upon reasonable request and with permission from the Instituto de Medicina Integral Prof. Frnando Figueira (IMIP) – www.imip.org.br

## Authors’ contributions

JGBA and FJC conceived the study. FJC, CAA, JRSJ, and MRT participated in the data collection. JGBA and JNF analyzed the data. JGBA and FJC wrote the main manuscript text. All authors reviewed the manuscript.

## Acknowledgments

The authors would like to thank to the CNPq (Brazilian National Council for Scientific and Technological Development), who financed this study, grant number 401907/2020-1.

